# In Parkinson’s disease, affective and chronic fatigue syndrome symptoms are associated with neuronal damage markers

**DOI:** 10.1101/2024.05.20.24307640

**Authors:** Hussein Kadhem Al-Hakeim, Hayder Naji Khudhair, Sayed-Omid Ranaei-Siadat, Fataneh Fatemi, Fateme Mirzajani, Mengqi Niu, Michael Maes

**Affiliations:** Department of Chemistry, College of Science, University of Kufa, Iraq.; Protein Research Center, Shahid Beheshti University, Tehran, Iran.; Sichuan Provincial Center for Mental Health, Sichuan Provincial People’s Hospital, School of Medicine, University of Electronic Science and Technology of China, Chengdu 610072, China; Key Laboratory of Psychosomatic Medicine, Chinese Academy of Medical Sciences, Chengdu, 610072, China; Department of Psychiatry, Faculty of Medicine, Chulalongkorn University, Bangkok, Thailand; Ph.D. Program in Mental Health, Department of Psychiatry, Faculty of Medicine, Chulalongkorn University, Bangkok, Thailand; Cognitive Impairment and Dementia Research Unit, Faculty of Medicine, Chulalongkorn University, Bangkok, Thailand; Cognitive Fitness and Biopsychological Technology Research Unit, Faculty of Medicine Chulalongkorn University, Bangkok, 10330, Thailand, Bangkok, 10330, Thailand; Department of Psychiatry, Medical University of Plovdiv, Plovdiv, Bulgaria; Research Institute, Medical University of Plovdiv, Plovdiv, Bulgaria; Kyung Hee University, 26 Kyungheedae-ro, Dongdaemun-gu, Seoul 02447, Korea; Research and Innovation Program for the Development of MU - PLOVDIV–(SRIPD-MUP)“, Creation of a network of research higher schools, National plan for recovery and sustainability, European Union – NextGenerationEU

**Keywords:** Chonic fatigue syndrome, neuroimmune, inflammation, mood disorders, depression, biomarkers

## Abstract

**Background:** Parkinson’s disease (PD) is frequently accompanied by mood and chronic fatigue syndrome (CFS) symptoms. It is unknown whether immune activation and insulin resistance (IR) or brain injuries impacts the severity of affective and CFS symptoms due to PD.

**Aims:** To examine whether immune, IR, and/or brain injury biomarkers determine affective and CFS symptoms due to PD.

**Methods:** Using a case (70 PD patients) control (60 healthy controls) study design, we assessed affective and CFS symptoms, measured the peripheral immune-inflammatory response system (IRS) using interleukin-6 (IL-6), IL-10, zinc, and calcium levels, the Homeostasis Model Assessment 2 insulin resistance (HOMA2IR) index, and serum brain injury markers including S100 calcium-binding protein B (S100B), neuron-specific enolase (NSE), phosphorylated tau217 (pTau217), and glial fibrillary acidic protein (GFAP).

**Results:** PD patients showed increased affective and CFS scores, IRS activation, HOMA2IR, NSE, GFAP, pTau217, and S100B levels as compared to controls. A large part (52.5%) of the variance in the mood+CFS score was explained by the regression on NSE, S100B, HOMA2IR index, interleukin-10 (IL-10) (all positively) and calcium (inversely). The HOMA2IR and IRS indices were significantly associated with all 4 brain injury biomarkers. A large part of the variance in the latter markers (37.0%) was explained by the cumulative effects of the IRS and HOMA2IR indices.

**Discussion:** RS activation and IR in patients with PD contribute to damage to glial cell projections and type III intermediate filament, which in turn contribute to affective and CFS symptoms.

## Introduction

Parkinson’s disease is a neurodegenerative disorder that affects a significant number of individuals aged 60 years and above. In fact, it is considered the most prevalent movement disorder globally (Uwishema et al., 2022). Parkinson’s disease is characterized by alterations in motor function, such as tremors, rigidity, bradykinesia, and postural instability (Saini et al., 2024). Additional symptoms that may arise include loss of smell, decline in cognitive function, alterations in personality, feelings of sadness, increased anxiety, persistent fatigue, and dysautonomic symptoms (Silva et al., 2023).

It has been observed that individuals diagnosed with Parkinson’s disease often experience affective symptoms, including depression and anxiety, as well as associated cognitive disorders (Chikatimalla et al., 2022; Marsh, 2013). Forty to fifty percent of individuals diagnosed with Parkinson’s disease experience symptoms of depression, with around 50% of them being diagnosed with major depression (MDD) (Marsh, 2013). Chronic fatigue is a common neuropsychiatric symptom complex in Parkinson’s disease that impacts over 50% of patients. It significantly reduces their quality of life and ability to conduct daily tasks (Siciliano et al., 2018). Chronic fatigue is one of the most distressing symptoms experienced by patients with Parkinson’s disease that is frequently reported in the literature (Müller et al., 2013).

Parkinson’s disease is a complex condition characterized by the degeneration of neurons and microglial activation (neuroinflammation) in the nervous system. It has been found that the activation of immune-inflammatory pathways in the peripheral system plays a significant role in this disorder (Chikatimalla et al., 2022). Both major depression and chronic fatigue syndrome (CFS) exhibit peripheral IRS activation, leading to microglial activation (Maes and Carvalho, 2018; Twisk and Maes, 2009). Elevated serum levels of proinflammatory cytokines, such as interleukin (IL)-6, and immunoregulatory cytokines, like IL-10, have been found in Parkinson’s disease, MDD, and chronic fatigue syndrome (CFS) (Maes and Carvalho, 2018; Morris and Maes, 2013; Qin et al., 2016).

Other potential pathways that contribute to the development of depression and CFS symptoms in Parkinson’s disease include damage to neurons, microglia, and astroglia. Cell death is a common consequence of cellular structure damage (Manev et al., 1989). As a result, there is an observed rise in the levels of neuronal or astroglial damage biomarkers in serum (Huibregtse et al., 2021; Silvestro et al., 2024; Wang et al., 2015). In cases of brain injury or neurodegenerative conditions, certain markers in the blood, such as S100 calcium-binding protein B (S100B), neuron-specific enolase (NSE), phosphorylated tau217 (pTau217), and glial fibrillary acidic protein (GFAP), have been found to be elevated (Brenner, 2014; das Neves et al., 2021; Palmqvist et al., 2020; Palumbo et al., 2008). Emerging research suggests that certain medical conditions such as Parkinson’s disease, MDD, and CFS may be associated with elevated levels of specific markers indicating neuronal, microglial, or astroglial damage. Several studies have reported on this phenomenon (Al-Hakeim et al., 2023b; Andersson et al., 2011; Dutta, 2021; Gonzales et al., 2021; Lin et al., 2023; Ridhaa et al., 2023; Rydbirk et al., 2017).

There is evidence from epidemiological studies suggesting a potential link between diabetes and an elevated risk of Parkinson’s disease, or the possibility that insulin resistance (IR) may play a role in the progression of the disease (Chohan et al., 2021; Liu and Tang, 2021). IR has been found to have a negative impact on the progression of Parkinson’s disease (Ruiz-Pozo et al., 2023), MDD (Morelli et al., 2021), and symptoms related to affective disorders and CFS caused by Long COVID (Al-Hakeim et al., 2023c; Vojdani et al., 2024).

However, it is currently unknown whether IRS activation and increased IR are associated with signs of brain injuries (elevated serum levels of S100B, NSE, pTau217, and GFAP) in Parkinson’s disease and whether elevated serum levels of these brain injury markers impact the severity of affective and CFS symptoms.

Hence, the current study has been conducted to examine whether serum levels of S100B, NSE, pTau217, and GFAP are associated with the affective and CFS symptoms due to Parkinson’s disease and whether activation of immune-inflammatory pathways and increased IR are associated with the biomarkers of brain injuries.

## 2. Subjects and Methods

### Participants

This case-control study examined a group of seventy patients diagnosed with Parkinson’s disease, as well as a control group of sixty individuals who were in good health. The patients were registered at the Middle Euphrates Center for Neurological Sciences, located in Najaf City, Iraq. The diagnosis of Parkinson’s disease in all patients was made based on the clinical diagnostic criteria from the UK Parkinson’s Disease Brain Bank, as determined by two experienced neurologists (Hughes et al., 1992; Perlmutter, 2009). The study did not include individuals with diabetes mellitus type 1 or other systemic autoimmune diseases, neurodegenerative or neuroinflammatory disorders other than Parkinson’s disease, multiple sclerosis, rheumatoid arthritis, inflammatory bowel disease, liver or renal disease, or any other medical conditions. Individuals with axis-1 disorders, such as bipolar disorder, anxiety disorder, schizophrenia, autism, and substance use disorders, were not eligible to participate in the study. Furthermore, this study did not include patients who had experienced any affective disorder before developing Parkinson’s disease. These disorders encompass MDD, dysthymia, chronic fatigue syndrome, fibromyalgia, and psychotic disorders like delirium.

Prior to their involvement in the study, both control and patient participants were required to give written consent. They were provided with detailed information beforehand. The study was granted approval by the institutional ethics committee of the University of Kufa (2109/2023). The study followed ethical and privacy laws both in Iraq and internationally. It complied with various guidelines and declarations, such as the International Conference on Harmonization of Good Clinical Practice, the Belmont Report, the CIOMS Guidelines, and the World Medical Association’s Declaration of Helsinki. In addition, our institutional review board adheres to the International Guidelines for Human Research Safety (ICH-GCP).

### Clinical measurements

An experienced neurologist conducted a semi-structured interview to evaluate and gather socio-demographic and clinical information from both control subjects and patients. An expert in the field of neurology evaluated the extent of the motor and non-motor symptoms associated with Parkinson’s disease by employing the Movement Disorders Society Revision of Unified Parkinson’s Disease Rating Scale (MDS-UPDRS) as outlined in the studies conducted by Goetz et al. in 2007 and 2008 (Goetz et al., 2007; Goetz et al., 2008). The MDS-UPDRS is divided into four parts; part I: non-motor Experiences of Daily Living (nM-EDL), part II: motor Experiences of Daily Living (m-EDL), part III: motor examination, and part IV: motor complications (Goetz et al., 2008). Neurologists conducted the MDS-UPDRS assessment on both patients and controls, utilizing the ratings from the 4 domains for statistical analysis.

In addition, we have calculated two subdomains using the nM-EDL subdomain: mood symptoms (MOOD) and chronic fatigue syndrome (CFS) symptoms. MOOD was formulated as the summed combination of depressed mood, anxious mood, apathy, sleep problems, and fatigue. The concept of CFS involves the summed combination of cognitive impairment, sleep problems, pain and other sensations, daytime sleepiness, constipation problems, lightheadedness, and fatigue. As a result, we calculated the total of several factors such as depressed mood, anxious mood, apathy, sleep problems, fatigue, cognitive impairment, pain and other sensations, daytime sleepiness, constipation problems, and light-headedness. This total sum is labeled as MOOD+CFS. By utilizing the summation of MOOD+CFS scores, the patients were categorized into two distinct groups, employing a cut-off value of 14.5. Consequently, the patients can be divided into two subgroups: one consisting of individuals with low MOOD+CFS scores (n=40, range: 4-14), and the other consisting of those with higher MOOD+CFS symptoms (n=30, range 15-32). Therefore, we divided the patients with Parkinson’s disease into two subgroups and conducted a comparison of clinical and biomarker measurements among three groups: normal controls, and the two subgroups of Parkinson’s patients. The BMI is determined by dividing the weight in kilograms by the square of the height in meters. Tobacco use disorder (TUD) was diagnosed using the DSM-5 criteria.

### Assays

A volume of 5 milliliters of fasting blood samples was collected at approximately 9:00 a.m. After a brief waiting period, the blood samples, which had formed clots, were placed in a centrifuge and spun at a speed of 1200 Xg for a duration of five minutes. Subsequently, the serum was carefully divided and distributed among three Eppendorf tubes. Excluded from the study were samples that had undergone hemolysis. The tubes were subsequently frozen at -80 °C and remained in this state until they were thawed for the assays.

We utilized sandwich ELISA techniques to quantify the levels of serum human GFAP, IL-6, IL-10, insulin, NSE, pTau217, and S100B. These measurements were conducted using pre-made ELISA kits provided by Nanjing Pars Biochem Co., Ltd. (Nanjing, China). The coefficient of variation (CV) for all ELISA kits was less than 10.0%. We employed sample dilutions for samples that contained analytes with elevated concentrations. The spectrophotometric measurement of serum glucose, calcium, magnesium, copper, and zinc was performed using kits provided by Spectrum Diagnostics Co., (Cairo, Egypt). After analyzing the blood samples, we have calculated two indices: the first is the activation of the immune-inflammatory response system (IRS), which involves the z transformation of IL-6 (z IL-6) + z IL-10, - z zinc - z calcium (labeled as Comp_IRS). The second index that was examined is glial projection toxicity, which is computed as z NSE + z S100B + z GFAP + z pTau (labeled as Comp_GPT). The Homeostasis Model Assessment 2 (HOMA2) calculator, developed by the Diabetes Trials Unit at the University of Oxford, is a valuable tool for assessing homeostasis (https://www.dtu.ox.ac.uk/homacalculator/download.php). The HOMA2%S and HOMA2IR calculations were employed to determine insulin sensitivity and IR, respectively, based on the fasting serum insulin and glucose levels.

### Statistical analysis

In order to examine the distribution types of the results group, the Kolmogorov-Smirnov test was implemented. The researchers employed analysis of variance (ANOVA) or the Kruskal-Wallis test (KWT) to assess variations in continuous variables across different groups. The researchers utilized protected pairwise post-hoc analyses in order to investigate the variations among group means. To examine associations between nominal variables, the χ^2^-test or Fisher exact probability test was utilized in conjunction with contingency tables. Utilizing Pearson’s product-moment correlation coefficients, the correlations between variables were examined. Using multivariate general linear model (GLM) analysis (followed by tests of between-subject effects), the associations between categories and biomarkers were investigated, accounting for confounding variables including age, sex, and body mass index (BMI). By employing manual and automatic stepwise multiple regression analyses with significance levels of biomarkers (p-to-enter was 0.05 and p-to-remove was 0.06), we identified the biomarkers that were statistically significant in predicting the scores on the neuropsychiatric rating scale. An analysis was conducted on the impacts of each biomarker in both the entire study population and the subset of patients diagnosed with Parkinson’s disease. The standardized beta coefficients for each significant explanatory variable were computed utilizing the t statistic with the precise value of p, in addition to the model F statistic and total variance explained (R^2^). The latter was utilized to estimate the effect size of the model. In this study, statistical significance was established at a p-value of 0.05 using two-tailed tests. IBM SPSS 28 for Windows was utilized to analyze the data. The primary statistical analysis is the multiple regression analysis with MOOD+CFS score as the dependent variable and the biomarkers as explanatory variables. G*Power 3.1.9.7 showed that the a priori estimated sample size was 96 given a power of 0.8, alpha=0.005, effect size = 0.142 (corresponding to 12.5% explained variance), and 5 predictors.

Protein-protein interaction (PPI) analysis was conducted using String (STRING: functional protein association networks; string-db.org), as previously described. The organism under consideration was Homo sapiens, and the minimal interaction score was 0.7. In our case, we exclusively used the initial shell, with no further interactions occurring.

## Results

### Sociodemographic and clinical data

**Table 1** contains the demographic and clinical information of Parkinson’s disease (PD) patients, categorized by the severity of their MOOD+CFS symptoms into high or low subgroups, in addition to the control subjects. Age, sex, employment status, rural-to-urban ratio, marital status, TUD, BMI, and years of education did not differ significantly between the study groups, according to the findings. In terms of age of onset and disease duration, there were no significant differences between the low and high MOOD+CFS subgroups. Patients who presented with high levels of MOOD+CFS symptoms exhibited higher EDL-Part III scores than those who had low MOOD+CFS scores. No substantial disparities were observed in the motor complications score or m-EDL between the two subgroups of patients.

**Table 1.** Demographic and clinical data in Parkinson’s disease (PD) patients divided into those with high levels of mood + chronic fatigue syndrome (CFS) symptoms (High MOOD+CFS) and those with lower MOOD+CFS symptoms, and healthy controls (HC).

| Variables | Controls <sup>A</sup><br>n=60 | Low MOOD+CFS <sup>B</sup><br>n=40 | High MOOD+CFS <sup>C</sup><br>n=30 | F/$\chi^2$ | df | p |
| --- | --- | --- | --- | --- | --- | --- |
| Age Yrs | 62.5±5.0 | 61.3±11.1 | 65.0±10.1 | 1.59 | 2/127 | 0.207 |
| Sex Female/Male | 19/41 | 15/25 | 9/21 | 0.54 | 2 | 0.765 |
| Employment No/Yes | 49/11 | 34/6 | 24/6 | 0.33 | 2 | 0.850 |
| Rural/Urban ratio | 19/41 | 11/29 | 12/18 | 1.25 | 2 | 0.536 |
| Single/Married | 1/59 | 1/39 | 0/30 | FET | - | 1.000 |
| Education Yrs | 9.4±4.2 | 8.7±4.7 | 9.7±3.6 | 0.62 | 2/127 | 0.542 |
| Duration of Disease Yrs | - | 4.58±3.07 | 4.40±4.06 | 0.05 | 1/68 | 0.829 |
| Age of onset Yrs | - | 57.0±11.5 | 62.2±12.3 | 3.35 | 1/68 | 0.072 |
| Family history No/Yes | 60/0 | 33/7 | 29/1 | FET | - | 0.001 |
| TUD No/Yes | 46/14 | 32/8 | 21/9 | 0.96 | 2 | 0.619 |
| BMI kg/m<sup>2</sup> | 27.69±4.64 | 28.35±3.30 | 27.46±3.16 | 0.52 | 2/127 | 0.598 |
| nM-EDL | 0 <sup>B,C</sup> | 10.95±2.94 <sup>A, C</sup> | 23.27±6.16 <sup>A, B</sup> | KWT | - | <0.001 |
| m-EDL | 0 <sup>B,C</sup> | 23.43±10.49 <sup>A</sup> | 24.37±9.59 <sup>A</sup> | KWT | - | <0.001 |
| Part III | 0 <sup>B,C</sup> | 60.25±24.94 <sup>A, C</sup> | 69.77±28.46 <sup>A, B</sup> | KWT | - | <0.001 |
| Part IV | 0 <sup>B,C</sup> | 11.78±6.49 <sup>A</sup> | 12.30±6.67 <sup>A</sup> | KWT | - | <0.001 |
| MOOD | 0 <sup>B,C</sup> | 6.88±1.73 <sup>A, C</sup> | 12.32±2.49 <sup>A, B</sup> | KWT | - | <0.001 |
| CFS | 0 <sup>B,C</sup> | 5.45±2.06 <sup>A, C</sup> | 12.37±3.73 <sup>A, B</sup> | KWT | - | <0.001 |
| MOOD+CFS | 0 <sup>B,C</sup> | 9.90±2.55 <sup>A, C</sup> | 19.97±4.41 <sup>A, B</sup> | KWT | - | <0.001 |

FET: Fisher exact test, KWT: Kruskal-Wallis test, BMI: body mass index, TUD: tobacco use disorder, m-EDL: Motor aspects of Experiences Daily Living, nM-EDL: Non-motor-EDL, Part III: motor examination score, Part IV: motor complications score. MOOD: mood symptoms of the nM-EDL; CFS: CFS symptoms of the nm-EDL; MOOD+CFS: sum of all MOOD and CSF symptoms.

Regarding Parkinson’s disease patients, no significant correlations were observed among the scores of EDL parts I, II, III, and IV. No statistically significant correlations were observed between the CFS scores and any of the MDS-UPDRS scores. A significant correlation was observed solely between the MOOD score and the MDS-UPDRS-Part III score (r=0.274, p=0.022, n=70).

### Differences in the biomarkers between PD patients’ groups and controls

The serum biomarkers of Parkinson’s disease patients, categorized by those with high or low MOOD+CFS symptoms, are presented in **Table 2**. A comparison with healthy controls is also provided. The findings indicated that the concentrations of magnesium, copper, zinc, and IL-6 did not differ significantly among the study groups. The levels of HOMA2IR, pTau217, and GFAP in both patient groups exhibited substantial increases when compared to the control group. Both patient groups exhibit reduced concentrations of calcium and HOMA2%S in comparison to the control group. There are significant differences in serum concentrations of NSE, S100B, and IL-10 among the three study groups, with concentrations rising from the control group to the low MOOD+CFS subgroup to the high MOOD-CFS subgroup. A notable distinction was observed among the three study groups in terms of the Comp_GPT and Comp_IRS indices, which exhibited an upward trend from the control group to the low and high MOOD-CFS groups.

**Table 2.** Serum biomarkers in Parkinson’s disease (PD) divided into those with high levels of mood + chronic fatigue symptoms (High MOOD+CFS) and those with lower MOOD+CFS symptoms (low MOOD+CFS), and healthy controls (HC).

| Parameters | HC <sup>A</sup> | Low MOOD+CFS <sup>B</sup> | High MOOD+CFS <sup>C</sup> | F | df | p |
| --- | --- | --- | --- | --- | --- | --- |
| Magnesium mM | 0.93 (0.02) | 0.98 (0.03) | 0.95 (0.03) | 1.28 | 2/123 | 0.282 |
| Calcium mM | 2.38 (0.03) <sup>B,C</sup> | 2.18 (0.03) <sup>A</sup> | 2.23 (0.04) <sup>A</sup> | 13.96 | 2/123 | <0.001 |
| HOMA2%S | 110.62 (6.74) <sup>B,C</sup> | 82.39 (8.34) <sup>A</sup> | 77.81 (9.64) <sup>A</sup> | 5.46 | 2/123 | 0.005 |
| HOMA2IR | 1.16 (0.09) <sup>B,C</sup> | 1.54 (0.12) <sup>A</sup> | 1.73 (0.13) <sup>A</sup> | 7.15 | 2/123 | 0.001 |
| Copper mg/l | 0.83 (0.04) | 0.91 (0.05) | 0.98 (0.06) | 2.11 | 2/123 | 0.126 |
| Zinc mg/l | 0.75 (0.03) | 0.77 (0.03) | 0.748 (0.04) | 0.08 | 2/123 | 0.923 |
| NSE ng/ml * | 0.66 (0.07) <sup>B,C</sup> | 0.91 (0.09) <sup>A,C</sup> | 1.50 (0.10) <sup>A,B</sup> | 33.52 | 2/123 | <0.001 |
| S100B pg/ml | 29.13 (1.95) <sup>B,C</sup> | 39.80 (2.42) <sup>A,C</sup> | 51.42 (2.79) <sup>A,B</sup> | 22.17 | 2/123 | <0.001 |
| pTau217 pg/ml * | 3.51 (0.27) <sup>B,C</sup> | 4.48 (0.34) <sup>A</sup> | 4.92 (0.39) <sup>A</sup> | 4.78 | 2/123 | 0.010 |
| GFAP pg/ml * | 3.15 (0.60) <sup>B,C</sup> | 5.87 (0.74) <sup>A</sup> | 5.26 (0.85) <sup>A</sup> | 6.07 | 2/123 | 0.003 |
| IL-6 pg/ml * | 3.85 (0.73) | 6.10 (0.90) | 5.83 (1.04) | 0.38 | 2/123 | 0.682 |
| IL-10 pg/ml * | 3.34 (0.62) <sup>B,C</sup> | 5.53 (0.77) <sup>A,C</sup> | 9.07 (0.89) <sup>A,B</sup> | 26.81 | 2/123 | <0.001 |
| Comp_GPT (z score) | -0.55 (0.51) <sup>B,C</sup> | 0.21 (1.01) <sup>A,C</sup> | 0.82 (1.08) <sup>A,B</sup> | 28.24 | 2/127 | <0.001 |
| Comp_IRS index (z score) | -0.96 (1.77) <sup>B,C</sup> | 0.50 (2.40) <sup>A,C</sup> | 1.26 (2.26) <sup>A,B</sup> | 12.83 | 2/127 | <0.001 |

\*: Processed in Log<sub>10</sub> transformation
HOMA2IR: Homeostatic Model Assessment of Insulin Resistance (HOMA2), NSE: neuron-specific enolase, GFAP: Glial fibrillary acidic protein, IL-6: interleukin-6, p-tau217: phosphorylated tubulin associated unit, S100B: S100 calcium-binding protein B, Comp\_GPT: a composite based on GFAP, NSE, S100B, and GFAP, reflecting glial cell projection damage; Comp\_IRS: a composite based on IL-6, IL-10, zinc and calcium reflecting immune activation.

### Correlations between physio-affective symptoms and biomarkers in all subjects

**Table 3** displays the intercorrelation matrix among serum biomarkers and the nM-EDL, MOOD, CFS, and MOOD+CSF scores for all study subjects (patients subgroups and controls). Correlations between all neuropsychiatric scores and HOMA2IR, NSE, S100B, pTau217, GFAP, and IL-10 are statistically significant. All neuropsychiatric disorders exhibited noteworthy inverse correlations with calcium and HOMA2%S.

**Table 3.** Intercorrelation matrix between biomarkers and the non-motor (nM) experiences of daily life (nM-EDL) subdomain.

| **Variables** | **Total nM-EDL** | **MOOD** | **CFS** | **MOOD+CSF** |
| --- | --- | --- | --- | --- |
| Magnesium | 0.079(0.372) | 0.067(0.450) | 0.102(0.250) | 0.085(0.339) |
| Calcium | -0.334(<0.001) | -0.383(<0.001) | -0.314(<0.001) | -0.344(<0.001) |
| HOMA2%S | -0.314(<0.001) | -0.289(0.001) | -0.326(<0.001) | -0.309(<0.001) |
| HOMA2IR | 0.374(<0.001) | 0.339(<0.001) | 0.382(<0.001) | 0.363(<0.001) |
| Copper | 0.084(0.341) | 0.118(0.182) | 0.078(0.378) | 0.087(0.325) |
| Zinc | -0.016(0.859) | 0.012(0.897) | -0.015(0.864) | -0.007(0.934) |
| NSE | 0.611(<0.001) | 0.536(<0.001) | 0.602(<0.001) | 0.583(<0.001) |
| S100B | 0.496(<0.001) | 0.474(<0.001) | 0.500(<0.001) | 0.495(<0.001) |
| pTau217 | 0.291(0.001) | 0.250(0.004) | 0.283(0.001) | 0.269(0.002) |
| GFAP | 0.275(0.002) | 0.239(0.006) | 0.295(0.001) | 0.271(0.002) |
| IL-6 | 0.081(0.362) | 0.041(0.645) | 0.100(0.258) | 0.070(0.431) |
| IL-10 | 0.576(<0.001) | 0.494(<0.001) | 0.581(<0.001) | 0.559(<0.001) |

HOMA2IR: Homeostatic Model Assessment of Insulin Resistance (HOMA2) insulin resistance, HOMA2%S: (HOMA2) insulin sensitivity, NSE: neuron-specific enolase, GFAP: Glial fibrillary acidic protein, IL-6: interleukin (IL)-6, pTau217: phosphorylated tubulin associated unit, S100B: S100 calcium-binding protein B, MOOD: mood symptoms of the nM-EDL, CFS: CFS symptoms of the nm-EDL, MOOD+CFS: sum of all MOOD and CSF symptoms.

The outcomes of the multiple regression analysis are presented in **Table 4**. The dependent variables in this analysis were the neuropsychiatric scores, while the explanatory variables were biomarkers. The analyses allowed for the effects of age, sex, education, TUD, and BMI. Regression #1 demonstrates that calcium (inversely associated), NSE, IL-10, and HOMA2IR (all three significantly and positively associated), accounted for 55.5% of the variance in the nM-EDL score. In the second regression analysis, calcium (inversely associated), S100B, IL-10, NSE, and HOMA2IR (all significantly and positively associated), accounted for 48.8% of the variance in the MOOD score. As shown in Regression No. 3, we determined that the regression on NSE, IL-10, HOMA2IR (which is positively associated), and calcium (which is inversely associated) could account for 54.8% of the variance in the CFS score. It is demonstrated in Regression #4 that the regression on NSE, IL-10, HOMA2IR, S100B (all positively correlated), and calcium (inversely associated) could account for 52.5% of the variance in the MOOD+CFS score. 31.0 percent of the variance in the MOOD+CFS score was accounted for by the regression on Comp_IRS (β=0.199, t=2.38, p=0.019) and Comp_GPT (β=0.457, t=5.46, p<0.001) (F=32.34, df=2/127, p<0.001).

**Table 4.** Results of multiple regression analysis with physio-affective symptom domains as dependent variables and biomarkers as explanatory variables in all subjects of the study.

| Dependent Variables | Explanatory Variables | $\beta$ | t | P | F model | df | p | R<sup>2</sup> |
| --- | --- | --- | --- | --- | --- | --- | --- | --- |
| **#1. Total nM-EDL** | **Model #1** |  |  |  | 38.97 | 4/125 | <0.001 | 0.555 |
|  | NSE | 0.352 | 4.93 | <0.001 |  |  |  |  |
|  | IL-10 | 0.326 | 4.66 | <0.001 |  |  |  |  |
|  | HOMA2IR | 0.217 | 3.52 | 0.001 |  |  |  |  |
|  | Calcium | -0.212 | -3.49 | 0.001 |  |  |  |  |
| **#2. MOOD** | **Model #2** |  |  |  | 23.61 | 5/124 | <0.001 | 0.488 |
|  | NSE | 0.255 | 3.19 | 0.002 |  |  |  |  |
|  | Calcium | -0.278 | -4.25 | <0.001 |  |  |  |  |
|  | S100B | 0.174 | 2.21 | 0.029 |  |  |  |  |
|  | IL-10 | 0.222 | 2.86 | 0.005 |  |  |  |  |
|  | HOMA2IR | 0.154 | 2.22 | 0.028 |  |  |  |  |
| **#3. CFS** | **Model #3** |  |  |  | 37.92 | 4/125 | <0.001 | 0.548 |
|  | NSE | 0.337 | 4.69 | <0.001 |  |  |  |  |
|  | IL-10 | 0.340 | 4.82 | <0.001 |  |  |  |  |
|  | HOMA2IR | 0.229 | 3.68 | <0.001 |  |  |  |  |
|  | Calcium | -0.192 | -3.13 | 0.002 |  |  |  |  |
| **#4. MOOD+CFS** | **Model #4** |  |  |  | 29.18 | 5/124 | <0.001 | 0.525 |
|  | NSE | 0.283 | 3.73 | <0.001 |  |  |  |  |
|  | IL-10 | 0.286 | 3.89 | <0.001 |  |  |  |  |
|  | Calcium | -0.226 | -3.65 | <0.001 |  |  |  |  |
|  | HOMA2IR | 0.173 | 2.63 | 0.010 |  |  |  |  |
|  | S100B | 0.152 | 2.05 | 0.043 |  |  |  |  |

HOMA2IR: Homeostatic Model Assessment of Insulin Resistance (HOMA2) insulin resistance, NSE: neuron-specific enolase, S100B: S100 calcium-binding protein B, IL: interleukin, nM-EDL: non-Motor aspects of Experiences Daily Living, MOOD: mood symptoms of the nM-EDL; CFS: chronic fatigue syndrome (CFS) symptoms of the nm-EDL; MOOD+CFS: sum of all MOOD and CSF symptoms.

### Correlation between physio-affective symptom and serum biomarkers in PD group

Correlations between biomarkers and clinical rating scales were calculated for the restricted patient population. There were significant correlations between the total nM-EDL score and the following variables: HOMA2IR (r=0.274, p=0.022, n=70), NSE (r=0.480, p<0.001), S100B (r=0.282, p=0.018), and IL-10 (r=0.492, p<0.001). The MOOD+CFS score demonstrated a significant association with HOMA2IR (r=0.252, p=0.036), NSE (r=0.408, p<0.001), S100B (r=0.272, p=0.023, and IL-10 (r=0.460, p<0.001).

The outcomes of various stepwise multiple regression analyses utilizing biomarkers as independent variables and clinical neuropsychiatric scores as dependent variables in the restricted patient subgroup are presented in **Table 5**. 32.7% of the variance in the nM-EDL score was accounted for by IL-10 and NSE (both positively associated), according to Regression #1. Regression #2 indicates that 9.0% of the variance in the MOOD score was accounted for by the model utilizing IL-10. IL-10 and NSE accounted for a substantial proportion of the variance (31.9%) in CFS (Regression #3) and the sum of MOOD+CSF (26.3%) in Regression #4. According to the findings of Regression #5, GFAP and HOMA2IR collectively accounted for a substantial proportion (26.3%) of the overall m-EDL variance.

**Table 5.** Results of multiple regression analysis with physio-affective symptom domains as dependent variables and biomarkers as explanatory variables in the restricted group of patients with Parkinson’s disease.

| **Dependent Variables** | **Explanatory Variables** | **$\beta$** | **t** | **P** | **F model** | **df** | **p** | **R<sup>2</sup>** |
| --- | --- | --- | --- | --- | --- | --- | --- | --- |
| **#1. Total nM-EDL** | **Model** |  |  |  | 16.25 | 2/67 | <0.001 | 0.327 |
|  | IL-10 | 0.346 | 3.09 | 0.003 |  |  |  |  |
|  | NSE | 0.326 | 2.91 | 0.005 |  |  |  |  |
| **#2. MOOD** | **Model** |  |  |  | 6.77 | 1/68 | 0.011 | 0.090 |
|  | IL-10 | 0.301 | 2.60 | 0.011 |  |  |  |  |
| **#3. CFS** | **Model** |  |  |  | 15.68 | 2/67 | <0.001 | 0.319 |
|  | IL-10 | 0.360 | 3.20 | 0.002 |  |  |  |  |
|  | NSE | 0.303 | 2.69 | 0.009 |  |  |  |  |
| **#4. MOOD+CSF** | **Model** |  |  |  | 11.93 | 2/67 | <0.001 | 0.263 |
|  | IL-10 | 0.347 | 2.96 | 0.004 |  |  |  |  |
|  | NSE | 0.253 | 2.16 | 0.035 |  |  |  |  |
| **#5. Total m-EDL** | **Model** |  |  |  | 11.95 | 2/67 | <0.001 | 0.263 |
|  | GFAP | 0.358 | 3.32 | 0.001 |  |  |  |  |
|  | HOMA2IR | 0.290 | 2.68 | 0.009 |  |  |  |  |

nM-EDL: non-Motor aspects of Experiences Daily Living, MOOD: mood symptoms of the nM-EDL; CFS: chronic fatigue syndrome (CFS) symptoms of the nm-EDL; MOOD+CFS: sum of all MOOD and CSF symptoms, m-EDL: Motor aspects of Experiences Daily Living, HOMA2IR: Homeostatic Model Assessment of Insulin Resistance (HOMA2) insulin resistance, NSE: neuron-specific enolase, GFAP: Glial fibrillary acidic protein, IL: interleukin.

### Associations between neuronal and immune markers

In the total study group, we found significant correlations between Comp_IRS and NSE (r=0.413, p<0.001, n=130), S100B (r=0.326, p<0.001), pTau217 (r=0.419, p<0.001), GFAP (r=0.311, p<0.001), and Comp_GPT (r=0.507, p<0.001). **Figure 1** shows the partial regression of NSE on Comp_IRS after adjusting for age and sex. The Comp_GPT index was significantly associated with IL-6 (r=0.497, p<0.001), IL-10 (r=0.526, p<0.001), HOMA2%S (r=-0.375, p<0.001), and HOMA2IR (r=0.405, p<0.001). The HOMA2IR was correlated with NSE (r=0.245, p=0.005), S100B (r=0.371, p<0.001), pTau217 (r=0.313, p<0.001), GFAP (r=0.246, p=0.005). We found that 37.0% of the variance (F=37.31, df=2/127, p<0.001) in Comp_GPT is explained by the combined effects of Comp_IRS (β=0.459, t=6.45, p<0.001) and HOMA2IR (β=0.340, t=4.78, p<0.001). **Figures 2 and 3** show the partial regressions of Comp_GPT on Comp_IRS and HOMA2IR, respectively.

**Figure 1.**
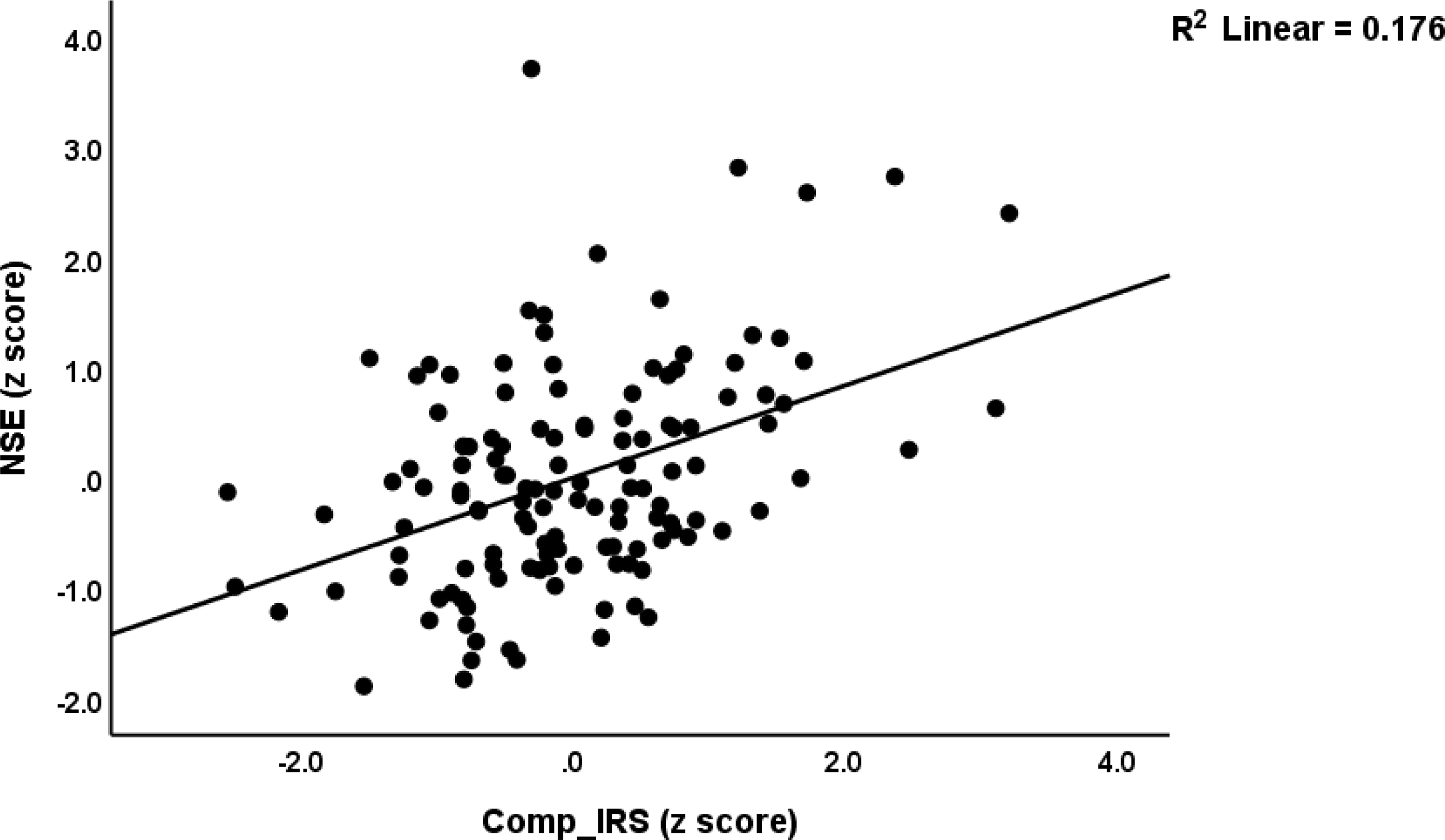
Partial regression of neuron-specific enolase (NSE) on a composite reflecting the immune-inflammatory response system (Comp_IRS) after adjusting for age and sex (p<0.001).

**Figure 2.**
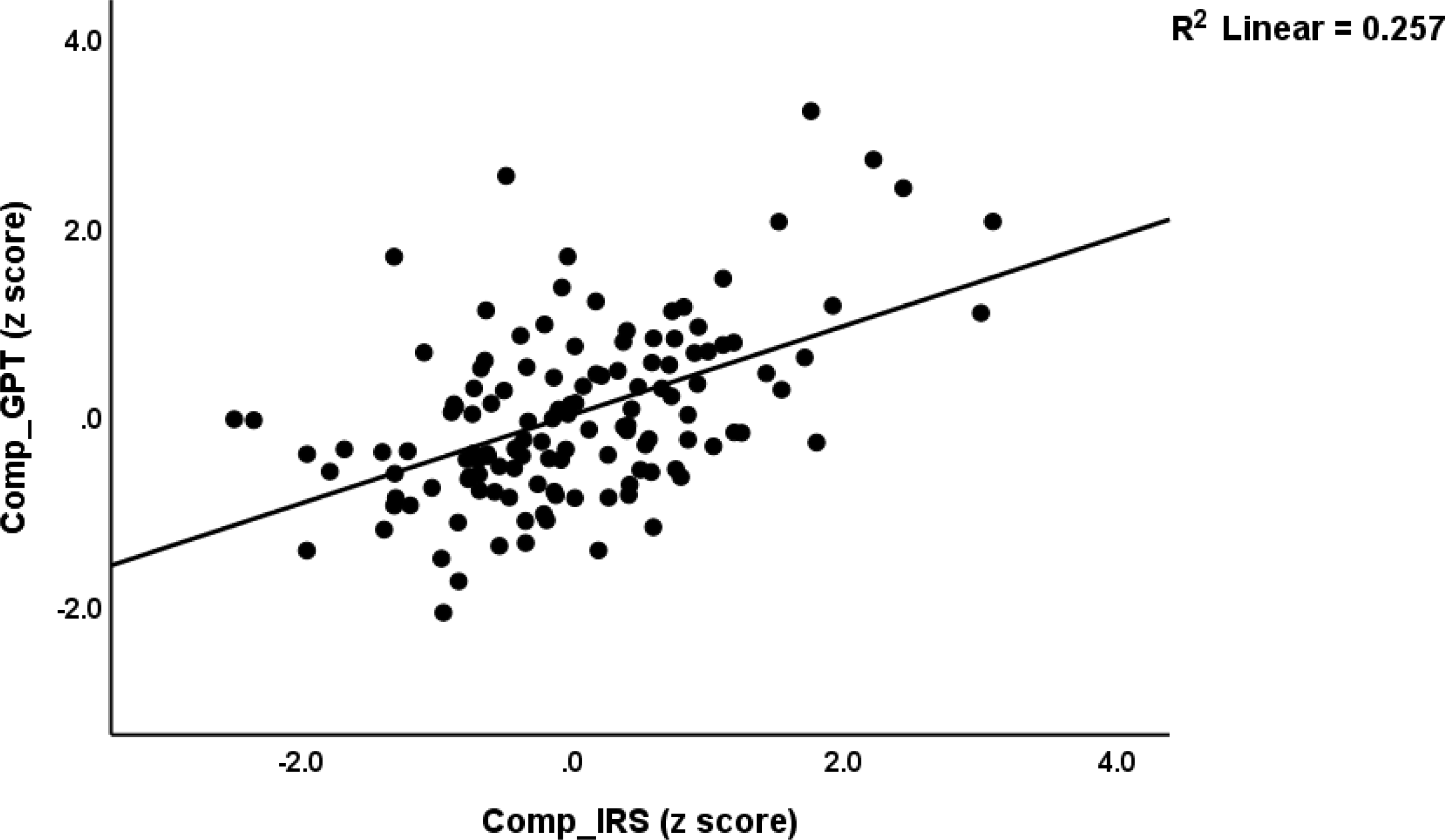
Partial regression of a composite score reflecting glial cell projection damage (Comp_GPT)

**Figure 3.**
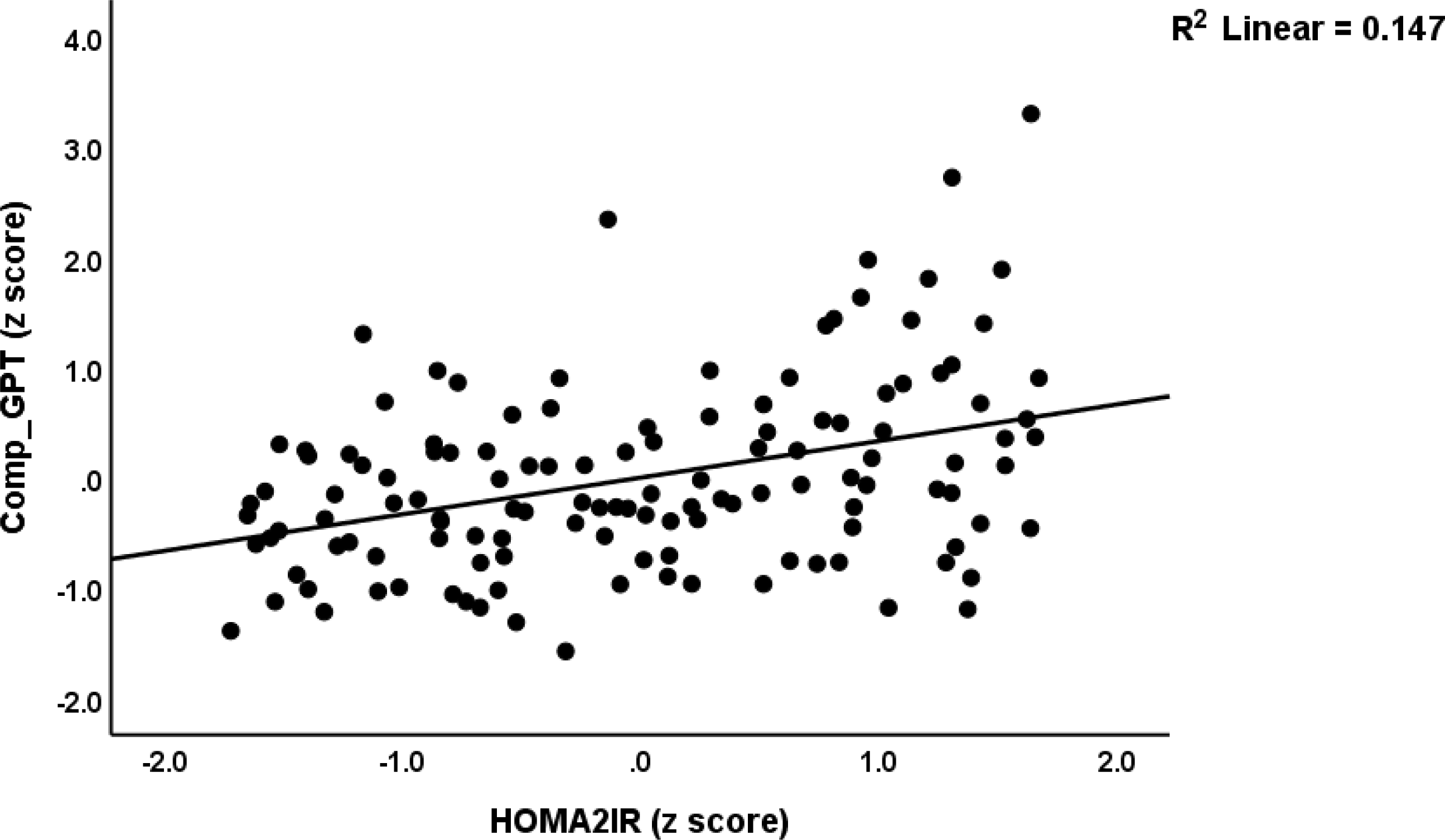
Partial regression of a composite score reflecting glial cell projection damage (Comp_GPT)

In the restricted patient group, we found significant correlations between Comp_IRS and NSE (r=0.319, p=0.007, n=70), S100B (r=0.264, p=0.027), pTau217 (r=0.432, p<0.001), GFAP (r=0.333, p=0.005), and Comp_GPT (r=0.467, p<0.001). In the patients, we found that the HOMA2IR was significantly correlated with NSE (r=0.289, p=0.015), S100B (r=0.377, p=0.001), pTau217 (r=0.343, p=0.004), GFAP (r=0.243, p=0.043). In patients, up to 36.3% of the variance in Comp_GPT (F=19.09, df=2/67, p<0.001) was explained by the regression on Comp_IRS (β=0.422, t=4.29, p<0.001) and HOMA2IR (β=0.383, t=3.90, p<0.001).

PPI analysis using String showed that these 4 genes shaped a tight network (number of nodes =4, edges=6, average clustering coefficient = 1, the expected number of edges =0; interaction score=0.7). This network was enriched in glial cell projections (FDR p=0.0246) and neuronal cell bodies (FDR p=0.0448), according to cellular component GO analysis. This network was found to be associated with neurofilament (FDR p=0.0047), type III intermediate filament (FDR p=0.0061), and polymeric cytoskeletal fiber (FDR p=0.0168), according to compartment subcellular analysis.

## Discussion

### Neuronal biomarker features of Parkison’s disease

The first major finding of this study is that the neuronal damage biomarkers NSE, S100B, pTau217, and GFAP are significantly elevated in Parkinson’s patients compared to the control group. Furthermore, these biomarkers are significantly associated with depression and CFS scores derived from the nM-EDL score. Moreover, NSE and S100B values were greater in Parkinson’s disease patients with elevated mood and CFS symptoms compared to those with decreased MOOD and CFS scores. In addition, significant correlations were observed between the MOOD+CFS scores and both NSE and S100B, even in the restricted study sample of Parkinson’s disease patients. These results indicate a correlation between the severity of neuronal damage markers and symptoms of MOOD and CFS, which may suggest that worsening neuronal damage in Parkinson’s patients could lead to the development of these conditions.

The most significant neuronal injury biomarker associated with MOOD and CFS symptoms in Parkinson’s disease is an increase in NSE. Increased NSE in the cerebro-spinal fluid has been suggested in some publications as a potential specific biomarker for Parkinson’s disease (Dutta, 2021). Neuronal damage (Constantinescu et al., 2009; Herrmann et al., 1999; Isgrò et al., 2015), neurodegenerative disorders such as stroke and Alzheimer’s disease (Chaves et al., 2010; Christl et al., 2019; Isgrò et al., 2015; Papuć and Rejdak, 2020), traumatic brain injury (Kim et al., 2018; Streitbürger et al., 2012), and hypoxic encephalopathy (León-Lozano et al., 2020) are all confirmed by elevated levels of NSE in serum and cerebrospinal fluid. Elevated NSE levels were found to be correlated with more severe depressive symptoms in children diagnosed with transfusion-dependent thalassemia (Dutta, 2021; Ridhaa et al., 2023). Conversely, adverse observations regarding serum NSE concentrations have been documented as well (Sathe et al., 2012; Schaf et al., 2005).

Previous research showed that patients with Parkinson’s disease exhibit a notable upregulation of S100B expression in brain tissues (Rydbirk et al., 2017) and a substantial increase in S100B levels in the substantia nigra (Sathe et al., 2012). Furthermore, a correlation has been observed between elevated nocturnal S100B levels and both sleep disruption and the severity of Parkinson’s disease (Carvalho et al., 2015). Blood samples from patients with Parkinson’s disease contained a greater quantity of antibodies against S100B than those from the control group (Wilhelm et al., 2007). An association between S100B polymorphisms and the age at which Parkinson’s disease manifests has been established in another study (Fardell et al., 2018). A systematic review found no significant correlations between the severity of depression and serum S100B levels, and no alterations in these levels were associated with major depression (Kozlowski et al., 2023).

S100B is a protein that is found in significant amounts in astroglial and oligodendroglial cells in the central nervous system. Consequently, the release of S100B by these cells could potentially indicate a glial reaction to inflammation, ischemia, and metabolic stress (das Neves et al., 2021). In addition, inflammation can disrupt the blood-brain barrier, which in turn triggers the activation of astrocytes. This activation results in an elevated release of S100B, as demonstrated in the study conducted by Zhang et al. in 2020 (Zhang et al., 2020). The levels of S100B in the serum are typically quite low. However, if these levels are elevated, it could suggest a disturbance in the blood-brain barrier and activation of astrocytes (Yu et al., 2020). It is worth noting that in stressful conditions, S100B has the potential to cause harmful effects by binding to receptors for advanced glycation end products (Donato et al., 2013). Furthermore, the presence of S100B triggers the activation of various molecules such as inducible nitric oxide synthase (iNOS), and proinflammatory cytokines including IL-6 and IL1β. This, in turn, leads to the development of oxidative stress and inflammation, as demonstrated by previous studies studies (Hu et al., 1997; Langeh and Singh, 2021). It is worth noting that decreased levels of S100B have been associated with potential benefits for the brain, such as supporting the survival of neurons (Bianchi et al., 2007), promoting neurogenesis (Baecker et al., 2020), and enhancing neuroplasticity (Nishiyama et al., 2002). These effects may contribute to improved memory and learning processes (Kleindienst et al., 2013).

A study conducted by Andersson et al. (2011) has found that the presence of Tau protein is a strong indicator of cognitive decline in individuals with Parkinson’s disease (Andersson et al., 2011). In the latter patients, the percentage of patients with an elevated t-tau/Aβ index increases from 15% to 29% when cognitive impairment is present, and this percentage further rises to 45% in patients with dementia caused by Parkinson’s disease (Montine et al., 2010). People who experience depression are more prone to early tau accumulation in specific areas of the brain that are important for managing emotions and forming memories (Gonzales et al., 2021). The pTau217 protein is widely observed in the microtubule system within cortical and hippocampal neurons, and to a lesser degree, in astrocytes and oligodendrocytes. The importance of tau proteins is found in their involvement in cell signaling, synaptic plasticity, and the maintenance of genomic stability (Guo et al., 2017). Plasma levels of this marker show an increase following neuronal damage, stress, or cognitive decline, as observed in studies by Luebke et al. (2023) (Luebke et al., 2023). and Green et al. (2023) (Green et al., 2023).

GFAP levels have been suggested as a potential biomarker for tracking the progression of Parkinson’s disease (Lin et al., 2023). Increased concentrations of GFAP and pTau217 have been found to be associated with MDD (Al-Hakeim et al., 2023a). Other authors have suggested that serum GFAP could serve as a potential marker for monitoring astroglial pathology in major depression (Steinacker et al., 2021). GFAP and its breakdown products are useful biomarkers for detecting and monitoring neurological conditions, such as Parkinson’s disease (Wang et al., 2018). Several studies have observed higher levels of GFAP in individuals with Parkinson’s disease when compared to healthy controls (Su et al., 2012). When astrocytes are injured, they release GFAP and its breakdown products into the interstitial/extracellular fluid (Lotankar et al., 2017; Luger et al., 2017).

Based on the findings, it can be concluded that Parkinson’s disease is characterized by the impairment of glial cell (astrocyte) projections, neurofilament, and specifically the type III intermediate filament. It appears that these impairments are closely linked to the development of affective and CFS symptoms in individuals with Parkinson’s disease. Previously, it was observed that MDD is accompanied by damage to neuronal and astroglial projections (Al-Hakeim et al., 2023a).

### Inflammation, insulin resistance and neuronal damage

Another significant discovery from this study reveals that the peripheral IRS activation in Parkinson’s disease patients is closely linked to heightened levels of all four glial cell damage markers and their composite score as well. Importantly, the detected associations were observed both in the combined group of controls and patients, as well as in the more specific subgroup of patients. These findings underscore the importance of the connections between peripheral inflammation and damage to glial projections and neurofilaments. Furthermore, both glial damage and IRS activation appear to play a significant role in explaining the variability in MOOD+CFS symptoms.

Activation of the peripheral IRS is believed to be involved in both Parkinson’s disease (Muñoz-Delgado et al., 2023) and MDD (Maes and Carvalho, 2018). In a previous study, it was demonstrated that there is a correlation between MDD and higher levels of GFAP and pTau217 (Al-Hakeim et al., 2023a). Parallel observations were made in cases of depression and CFS resulting from other medical conditions. For instance, in end-stage renal disease, the presence of affective and CFS-like symptoms is predicted by immune-inflammatory markers such as CRP and IL-10, as well as neuron damage biomarkers like serum neurofilament light chain protein, myelin basic protein, and nestin (Al-Hakeim et al., 2024). According to a study conducted by Ridhaa et al. (2023), the severity of affective and CFS-like symptoms in children with transfusion-dependent thalassemia can be predicted by assessing IRS activation (measured through IL-6, IL-10, CRP, zinc, and calcium levels) and neuronal damage (measured through serum NSE, GFAP, neurofilament light chain protein) (Ridhaa et al., 2023). In Long COVID, both immune activation and associated oxidative stress and increasing neuronal damage (as assessed with autoimmune response to myelin basic protein, synapsin, tubulin, myelin oligodendrocyte glycoprotein, and neurofilament chain protein) are associated with affective and CFS symptoms (Almulla et al., 2023).

Our findings suggest that IR plays a role in the development of affective and CFS symptoms in individuals with Parkinson’s disease, as well as the damage to glial projections. Indeed, we found that the HOMA2IR index has a significant impact on the physio-affective phenome, going beyond the effects of IRS activation and the glial damage biomarkers. Thus, a significant portion of the variability in the MOOD+CFS score could be accounted for by the regression analysis involving HOMA2IR, an IRS biomarker (specifically increased IL-10), and two markers related to brain injury (S100B and NSE). Recent research suggests a noteworthy link between heightened insulin resitance and Parkinson’s disease, indicating that insulin resistance could potentially enhance the progression of the condition (Parkinson’s Foundation, 2024). Recent research indicates that disrupted insulin signaling may have a negative impact on the advancement of neurodegeneration linked to Parkinson’s disease (Nowell et al., 2023). There are multiple mechanistic explanations for this correlation. One possibility is that insulin plays a protective role for dopaminergic neurons in the substantia nigra, shielding them from toxicity (Pang et al., 2016). Another explanation is that impaired insulin signaling disrupts dopamine homeostasis (Bassil et al., 2022; Gruber et al., 2023). Additionally, elevated blood sugar levels (hyperglycemia) may negatively impact dopaminergic neurons, leading to their programmed cell death through oxidative damage (Renaud et al., 2014; Su et al., 2020).

Patients exhibiting depressive symptoms may show signs of increased IR, as suggested by a study conducted by Morelli et al. in 2021 (Morelli et al., 2021). IR plays a crucial role in the development of affective and CFS symptoms associated with major depression (Al-Hakeim et al., 2023a), Long COVID (Al-Hakeim et al., 2023c; Vojdani et al., 2024), type-2 diabetes mellitus (Al-Hakeim et al., 2022), and atherosclerosis (Mousa et al., 2022), going beyond the impact of IRS activation or brain injury biomarkers. Several studies have found associations between fatigue scores in Parkinson’s disease and various biomarkers, including inflammatory and anti-inflammatory markers (Herlofson et al., 2018; Wang et al., 2021), IR (Özer et al., 2019), and biomarkers indicating neuronal damage (De Dreu et al., 2020; Wang et al., 2021).

#### Limitations

This study would have been more interesting if we had measured immune profiles based on cytokine/chemokines/growth factor assessment using a multiplex assay.

#### Summary

Compared to controls, Parkinson’s disease patients have higher levels of S100B, GFAP, HOMA2IR, NSE, and pTau217 indicating damage to glial cell projection and neurofilament. Moreover, Parkinson’s disease is characterized by peripheral IRS activation, a significant increase in IR, and increased affective and CFS symptoms. NSE, SB100, HOMA2IR index, interleukin-10 (IL-10) (all positively) and calcium (inversely) accounted for a significant portion (52.5%) of the variance in the affective + CFS scores. All brain injury biomarkers measured in the current study showed a strong correlation with the HOMA2IR and IRS indices. The combined impacts of the HOMA2IR and IRS indices accounted for a significant portion of the variance in the later markers (37.0%). Affective and CFS symptoms due to Parkinson’s disease are a result of damage to glial cell projections and type III intermediate filament. The latter may be caused by IRS activation and IR in PD patients. It follows that IRS activation and IR are new drug targets to treat affective and CFS symptoms due to Parkinson’s disease. This treatment should be augmented by treatments targeting the damage of glial cells with neurotrophic compounds.

#### Ethics statement

The study has been approved by the institutional ethics committee of the University of Kufa (2109/2023). The study adhered to both Iraqi and international ethical and privacy laws, including the International Conference on Harmonization of Good Clinical Practice, the Belmont Report, the CIOMS Guidelines, and the World Medical Association’s Declaration of Helsinki.

#### Consent for publication

Before participating in this study, each subject provided written informed consent.

## Funding

There was no funding for this study.

## Conflict of interest

There is no financial or other conflict of interest between the authors and any organization associated with the submitted paper.

## Author’s contributions

All the contributing authors have participated in the preparation of the manuscript.

## Data availability statement

The database created during this investigation will be provided by the corresponding author (MM) upon a reasonable request once the authors have thoroughly used the data set.

## Acknowledgments

We acknowledge the assistance of the workers at the Middle Euphrates Center for Neurological Sciences, Najaf City, Iraq in the sample collection and lab measurements.

## Notes

### Competing Interest Statement

The authors have declared no competing interest.

